# Childhood Adversity and Adolescent Epigenetic Age Acceleration: The Role of Adolescent Sleep Health

**DOI:** 10.1101/2024.09.02.24312939

**Authors:** Karissa DiMarzio, Darlynn M. Rojo-Wissar, Evelyn Hernandez Valencia, Mikayla Ver Pault, Shane Denherder, Adamari Lopez, Jena Lerch, Georgette Metrailer, Sarah M. Merrill, April Highlander, Justin Parent

**Author notes:** **Corresponding Author:** Justin Parent, PhD, University of Rhode Island, 142 Flagg Road, Kingston, RI 02881.

## Abstract

**Study Objectives:** We investigated how a dimension of early life adversity (ELA) capturing threat in the home relates to later epigenetic age acceleration in adolescence through sleep (duration, efficiency, and timing), to empirically test theoretical models suggesting the importance of sleep as a key mechanism linking ELA with poor health outcomes, and to expand the limited literature on sleep and epigenetic aging among youth.

**Methods:** We utilized data from 861 participants from the Future of Families and Child Wellbeing Study (FFCWS) who participated in the actigraphy sub study at age 15. Sleep variables used were average total sleep time (TST), sleep efficiency (SE), and sleep onset timing. Home threat was determined at ages 3, 5, and 9 from parent reports on the Child Conflict Tactics Scale (CTS-PC), and epigenetic aging was measured through DNA methylation analyses of saliva samples collected at age 15.

**Results:** Higher levels of childhood home threat exposure were associated with less adolescent TST, lower SE, and later sleep onset timing. Adolescent SE and timing were associated with a faster pace of aging and epigenetic age acceleration. Sleep efficiency and timing mediated the link between childhood home threat exposure and adolescent epigenetic aging.

**Conclusions:** Epigenetic embedding of childhood threat exposure in the home may occur through adversity-related sleep disturbances in adolescence. Findings warrant greater attention to pediatric sleep health in theoretical models of biological embedding of adversity and point to the examination of improving sleep health as a potential way to prevent adversity-related epigenetic age acceleration.

**Statement of Significance:** This study brings together two largely disparate bodies of literature on early life adversity and epigenetics and sleep and epigenetics. Findings advance knowledge on how adversity impacts specific aspects of sleep (duration, efficiency, and timing), and biological aging, offering potential pathways for mitigating long-term health risks associated with childhood adversity.

---

Early life adversity, which has been conceptualized in many different ways, is common and associated with poor outcomes across the lifespan. For example, according to a recent meta-analysis (Madigan et al., 2023),^1^ half of the adult population in the United States (US) report experiencing at least one adverse childhood experience (ACE), and 16% report four or more. ACEs include experiences of child abuse and neglect as well as broader household dysfunction, like substance use problems and parental incarceration. Since the original ACE study^2^, research has consistently shown robust associations between ACEs and a wide range of negative physical and mental health outcomes, including many major causes of death.^3^ These findings underscore early adversity as a major public health concern and priority.^4^

One of the reasons that early life adversity is linked with these poor long-term health outcomes is that the physiological stress caused by early adversity can be so pervasive that it becomes biologically embedded.^5^ This occurs when responses to stress alter biological functions, which can happen across multiple systems, and lead to long-lasting changes in physiology. Biological embedding via epigenetic changes is one promising molecular mechanism involved in stress physiology and downstream health consequences.^6^ Briefly, epigenetic modifications to the genome allow for altered gene expression and chromatin conformation, but do not change the DNA sequence and thus permit elaboration of the genome beyond what is determined by DNA base coding. The most highly studied and best characterized epigenetic modification in humans, DNA methylation (DNAm), primarily involves a direct covalent, chemical modification of a cytosine base lying sequentially adjacent to a guanine base (CpG) which may associate with subsequent gene transcription.^7^

An area of recent substantial growth in biological embedding and epigenetics is the use of epigenetic “clocks” based on DNAm of Cpg sites that closely track biological aging processes.^8,9^ Epigenetic clocks have emerged as accurate estimators of aging in healthy children and adults. Aging faster epigenetically than expected chronologically is known as epigenetic age acceleration. Increased epigenetic age acceleration shows critical associations with early life adversity and, in turn, poor health outcomes.^9^ Hogan et al.^10^ found that early adversity characterized by home threat (e.g., physical or emotional abuse) predicted epigenetic age acceleration across adolescence, which, in turn, was associated with higher levels of psychopathology in the Future of Families and Child Wellbeing Study (FFCWS). Similarly, Chang and colleagues^11^ also found that childhood physical and emotional aggression in the home setting was associated with accelerated epigenetic aging in the FFCWS. A greater understanding of the mechanisms underlying associations between early life adversity and epigenetic aging would provide targets for intervention that could help mitigate poor health outcomes and improve long-term trajectories among individuals exposed to adversity. A mechanism that could link adversity to epigenetic changes is stress-related disruption to pediatric sleep health.

## Early Life Adversity and Sleep

Researchers have estimated that approximately half of all children in the US experience sleep problems, with this number growing as high as 70% during the transition to adolescence, which is characterized by physiological (e.g., delayed circadian phase) and socio-contextual (e.g., school start times) developmental shifts that affect sleep-wake regulation.^12–16^ For example, adolescents’ biological shift to later sleep timing paired with early school start times can increase their risk for insufficient sleep and circadian misalignment. Experiences of adversity likely compound existing risk for sleep and circadian disruption among adolescents, as a growing body of literature demonstrates that youth who have experienced adversity have a heightened risk for impaired sleep. Indeed, previous work using the FFCWS it was found that ACEs occurring earlier in childhood were associated with greater social jetlag, longer total sleep time during weekends (potentially indicating greater need for stress recovery), and greater insomnia symptoms at age 15;^17^ however, findings were limited to self-report measures and did not include objective assessments of sleep behaviors (e.g., actigraphy) — limitations that are consistent in the broader literature.^18,19^

Sleep disturbances, like with early adversity, are associated with poor health outcomes across the lifespan, and both appear to affect health via many of the same neurobiological mechanisms (e.g., disrupted regulation systems, hyperarousal).^20^ For example, research across rodent models and human youth and adults has shown that sleep deficiencies can result in increased pro-inflammatory cytokines and oxidative stress, and cortisol circadian rhythm disruption.^21,22,23,24^ Importantly, these systems are all vital to stress regulation, are implicated in the pathogenesis of stress-related psychiatric disorders^25^ and are integrally intertwined with chronobiology (e.g., circadian rhythms).^22,26^ As a result of these connections between early life adversity and sleep, and with each of those and health, sleep disturbances are proposed to be an important mechanism connecting early experiences of adversity to poorer health outcomes in the long term.^20^ Despite this evidence, the two bodies of literature on early adversity and epigenetics and on sleep and epigenetics remain comparatively disjointed, requiring greater exploration.

## Sleep and Epigenetic Aging

Like with early life adversity, there is evidence for poorer sleep health having epigenetic consequences — although this area has received less attention to date.^27^ Across species, there is growing evidence that sleep disturbances are tied to epigenetic modifications to the genome.^28^ Among human adults, for example, previous studies have found associations between insomnia symptoms, shorter sleep duration, and poorer sleep efficiency and epigenetic aging, particularly pace of aging and age acceleration.^29,30^ However, research on the connection between sleep and DNAm across childhood and adolescence is limited and mixed. For example, Koopman-Verhoeff et al.^31^ found that DNAm patterns were associated with shorter actigraphy-based sleep duration in childhood. However, this relationship was not observed with subjectively assessed sleep, which is consistent with other studies that have failed to find links between subjective assessments of sleep and epigenetic aging among youth.^32^ These findings underscore the importance of including objective sleep measurement tools in studies involving epigenetic aging. One recent study found that objectively assessed sleep initiation in adolescents was associated with altered young adult DNAm in genes previously identified in adult Genome-Wide Association Studies of sleep and circadian phenotype.^33^Only one study has examined the association between objectively assessed sleep and epigenetic aging in adolescence. Banker and colleagues^34,35^ used a cluster analysis method that included physical activity to provide initial evidence for an association between later sleep timing and accelerated epigenetic aging.

## The Current Study

Despite growing evidence that pediatric sleep health is disrupted by early adversity and that both adversity and sleep deficiencies are biologically embedded via epigenetic changes, particularly epigenetic age acceleration, no previous investigations have connected these two research areas. In the current study, we used data from the Future of Families and Child Wellbeing Study (FFCWS) to evaluate three domains of actigraphy-measured sleep (i.e., sleep duration, efficiency, and timing) as mediators of the association between early life adversity (captured as threat in the home), and epigenetic age acceleration. This builds on prior work in the FFCWS showing links between early life adversity and epigenetic age acceleration,^10^ and between early life adversity and subjective measures of sleep.^17^Further, it provides an empirical evaluation of theoretical models suggesting that sleep is a crucial mechanism linking early life adversity to poorer health outcomes,^20^f and expands the nascent sleep–epigenetics literature, particularly as it relates to youth.

## Methods

### Participants

Data for the current study are from the Future of Families and Child Wellbeing Study (FFCWS)-a prospective longitudinal birth-cohort study^36^ of 4,898 children born between 1998 and 2000 in 20 large U.S. cities. Participating families were recruited at child’s birth (1998-2000) and included married and non-married parents, with non-marital births oversampled at a rate of 3:1. Families were surveyed at approximately the child’s birth and ages 1, 3, 5, 9, 15, and 22 years. In the present study, we examined a subset of FFCWS participants (n = 861), who participated in the actigraphy sub-study and had at least two days of valid actigraphy data at age 15. Table 1 includes complete social demographic details of the FFCWS subsample using in the current study.

**Table 1.**
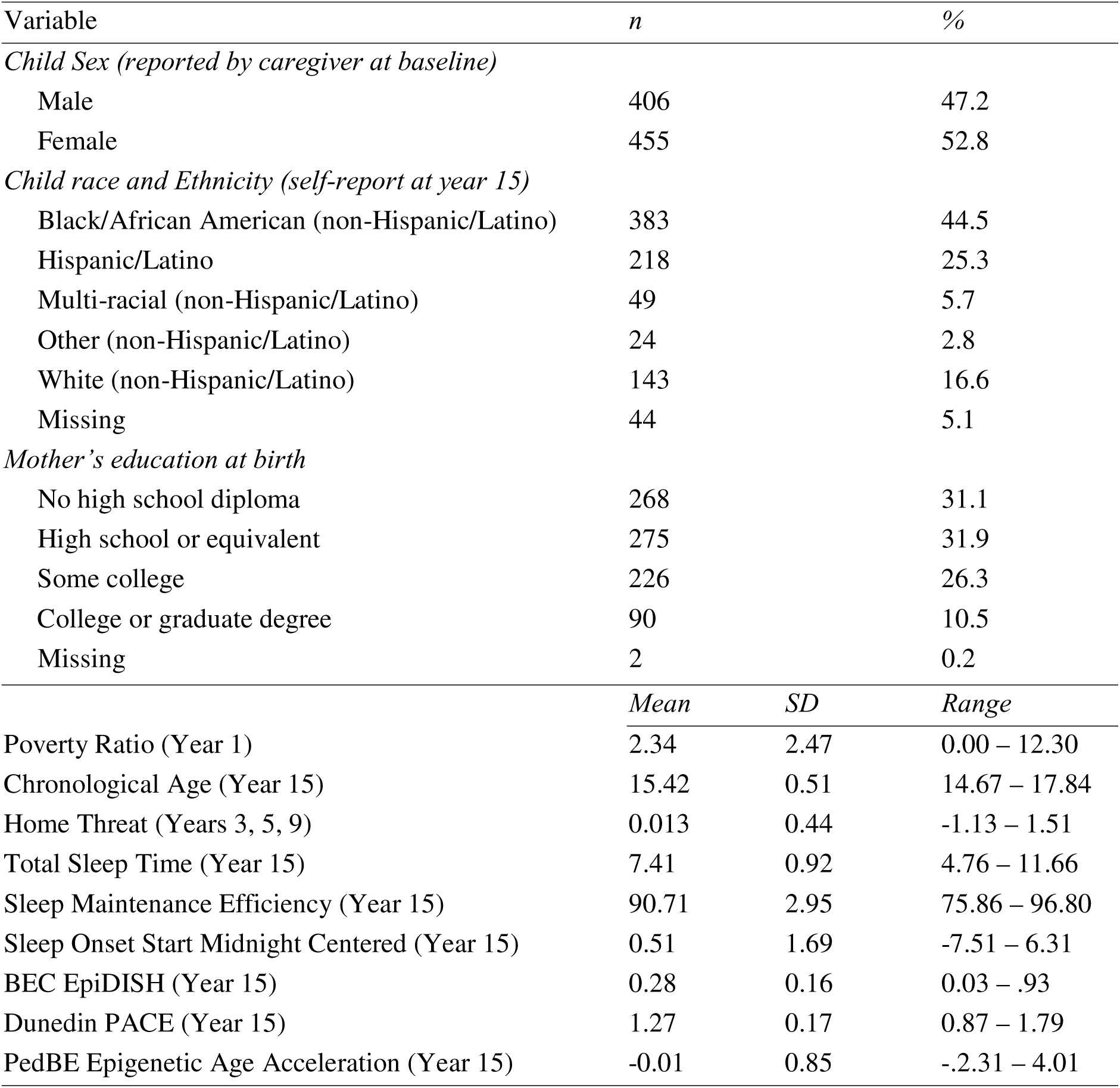
Descriptive Statistics of Study Participants.

### Measures

#### Home Threat

A latent variable was created to represent home threat at ages 3, 5, and 9 using the Parent-Child Conflict Tactics Scale (CTS-PC) based on previous studies using FFCWS data.^10,37^ Home threat was defined as being exposed to physical and/or emotional abuse by a primary caregiver. Occurrences of physical (“hit child on bottom with a hard object”) and emotional (“said you would send child away or would kick child out of the house”) abuse within the last year were reported by primary caregivers. Three items per category were included in the measure, and each item was rated on a 7-point Likert scale ranging from “0 - never happened” to “6 - more than 20 times”. The factor score of a global childhood home threat factor was extracted, representing home threat experiences across ages 3, 5, and 9, using similar items across waves. See Hogan et al. for complete details.

#### Sleep

Sleep variables were derived from wrist actigraphy devices (Actiwatch Spectrum; Philips-Respironics, Murrysville, PA) worn on the non-dominant hand for one week. Staff at the Sleep, Health, and Society Collaboratory at Penn State retrieved actigraphy data in 30-second epochs using Philips Actiware software version 6.0.4. Multiple trained scorers determined cut-points for sleep and validity of days following a validated algorithm.^38^ See Mathew et al.^39^ for complete details on scoring procedures. We used the following sleep variables computed by the Sleep, Health, and Society Collaboratory and made publicly available: Total sleep time, sleep maintenance efficiency, and sleep onset timing.

##### Total Sleep Time (TST)

The average total number of minutes of sleep a participant experienced in a 24-hour cut-point day, including nighttime sleep and any naps. This was converted to an hour metric to improve model estimation.

##### Sleep Maintenance Efficiency

The percentage of time spent asleep between nighttime sleep onset and offset. Higher values are indicative of better sleep quality (Nye & Buxton, 2022).

##### Sleep Onset Timing

Nighttime sleep onset time, defined as the last 30-second epoch of >10 activity counts followed by at least 5 consecutive epochs of activity counts ≤ 10. Sleep onset time is midnight centered, with values of 0 equaling midnight.

#### Epigenetic Aging

The FFCWS survey subcontractor, Westat Inc. arranged the sample collection. Westat interviewers used the Oragene® DNA Self-Collection kits (OGR-500) (DNA Genotek Inc) to collect child saliva samples during in-home visits for children at age 9 and 15. Available samples [n = 3,945] were assayed using methylation arrays (Infinium Human Methylation 450K and Infinium Methylation EPIC; Illumina) according to the manufacturer’s protocol. Samples were excluded if the ENmix R package quality control procedure identified samples as having outlier methylation or bisulfite conversion values or, if the sex predicted from the methylation data differed from the recorded sex. Two DNA methylation-based methods were used to estimate epigenetic aging.

##### DunedinPACE

The DunedinPACE pace of aging,^40^ previously employed in pediatric saliva samples,^41,42^ was examined for epigentic pace of aging. A value of one in this measure indicates the estimated epigenetic age and chronological age were equivalent, with a greater value indicating epigenetic age acceleration in comparison to chronological age. The development of this DNA methylation biomarker differs from age-focused epigenetic estimators in that it is rooted in the dynamics of health and phenotypes supporting successful biological aging processes, ^40,43^ including being trained on within-individual decline in 19 indicators of organ-system integrity across spanning two decades in the Dunedin Study.^40^ The FFCWS has two DunedinPACE measures, and the current study used the most updated method (poam45).

##### PedBE Epigenetic Age Acceleration

Analyses estimating DNA methylation age acceleration in children were conducted using the Pediatric Buccal Epigenetic Clock (PedBE),^44^ a clock trained in oral tissue to estimate biological age in children within an error of less than 4 months using 95 sites across the epigenome. PedBE was measured by the residuals of a linear mixed effect model with maximum likelihood estimation of predicted PedBE age on reported chronological age, accounting for predicted buccal epithelial cell proportion (as recommended by the authors of the tool)^44^ and a random effect of individual (both year 9 and 15 were included). This was completed in R (4.3.1) with the nlme package. Buccal epithelial cell proportion was estimated using the EpiDISH^45^ package and accounted for during epigenetic age acceleration calculation due to the association of this cell type and age.

##### Demographic Characteristics

At the child’s birth, the mother reported her own education level, household income and size, which were used to calculate their poverty ratio, and sex of the focal child. The focal child self-reported racial identity at age 15 years.

### Statistical Analysis Plan

A path analysis model was conducted using Mplus^46^, applying Full Information Maximum Likelihood (FIML) for any missing data, and maximum likelihood estimation with robust standard errors (MLR) was used to adjust for possible non-normality. The latent factor score of home threat across ages 3, 5, and 9 was the primary predictor in models. Year 15 Total sleep time, sleep maintenance efficiency, and sleep onset timing served as the simultaneous mediators. Year 15 Dunedin pace of aging and PedBE epigenetic age acceleration served as simultaneous outcome variables. Youth sex, Year 1 family poverty ratio, and buccal epithelial proportion were included as covariates. Youth self-report race and ethnicity and Year 9 epigenetic variables were included as auxiliary variables. The model indirect command in Mplus was used to obtain bootstrap standard errors for indirect effects. Sensitivity analyses examined the robustness of findings to summer-based actigraphy assessment, nighttime sleep not including naps instead of total 24-hour sleep time, the number of valid actigraphy days, quadratic effects, and multiple testing correction.

## Results

### Preliminary Analyses

Table 1 and Figure 1 depict the distribution of the primary study variables. On average, adolescents had insufficient sleep duration, adequate sleep efficiency (not including sleep onset latency), and sleep onset timing around midnight. Further, adolescents demonstrated an accelerated epigenetic pace of aging and an age-consistent PedBE epigenetic clock. However, substantial variability was present across all primary variables with sleep health parameters ranging from severe deficiencies to healthy levels and epigenetic aging ranging from decelerated to accelerated epigenetic aging. The correlation heatmap of core study variables is depicted in Figure 2. Bivariate associations were generally in the expected direction and supported primary models.

**Figure 1.**
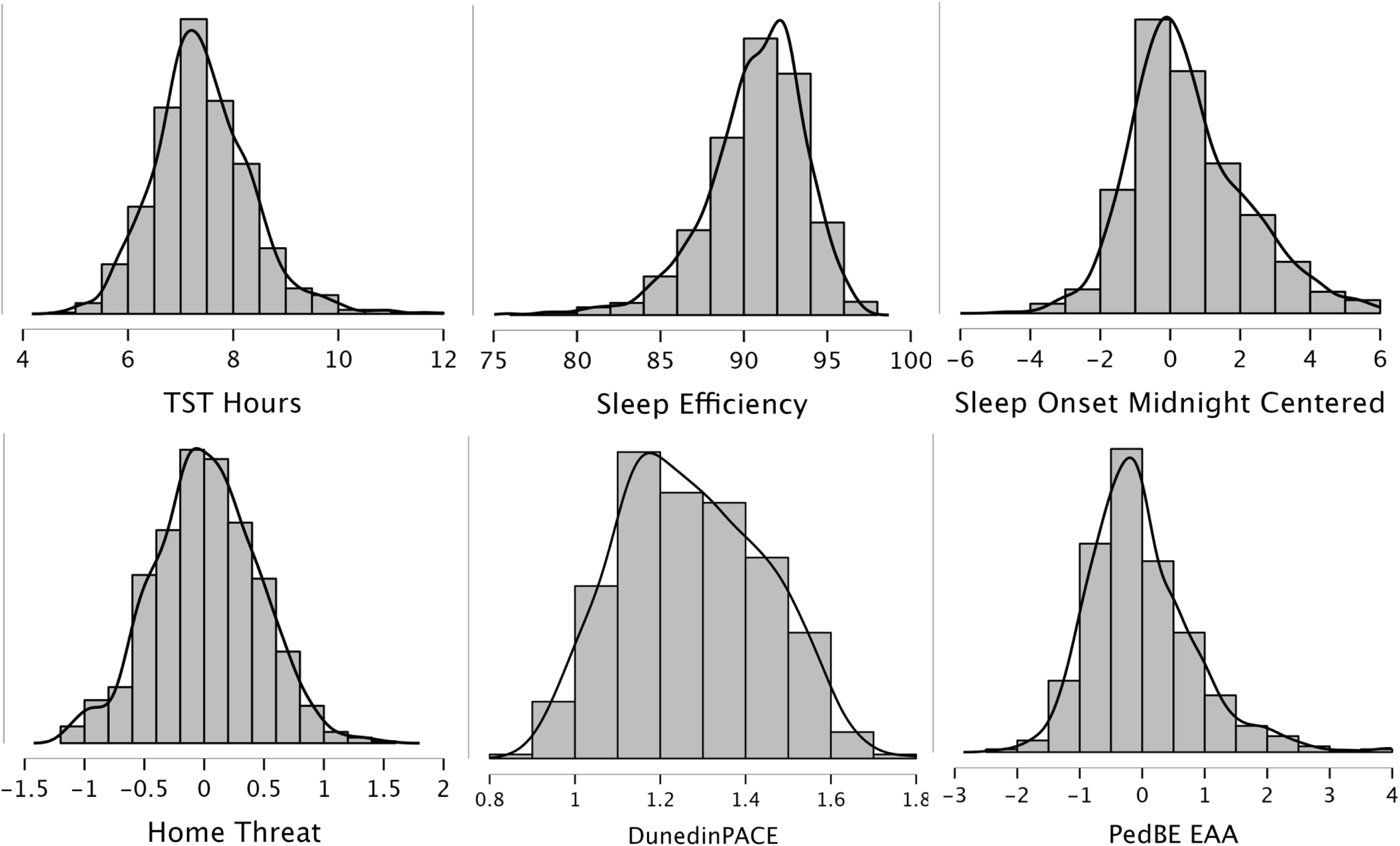
Density Histogram Plots of Primary Study Variables.

**Figure 2.**
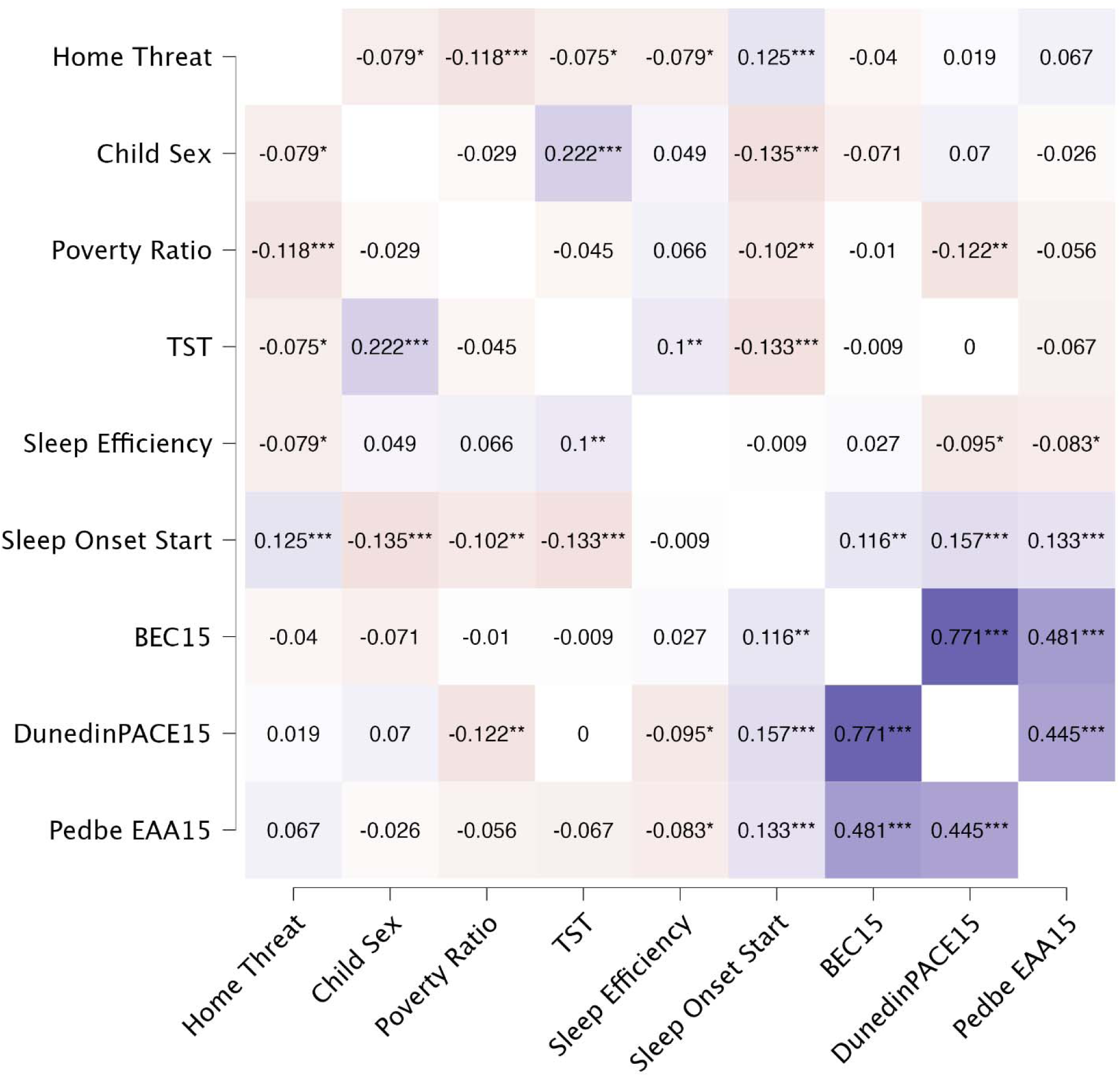
Heatmap Correlation Matrix of Primary Study Variables.

### Primary Analyses

Complete results are reported in Table 2, and a simplified mediation model is depicted in Figure 3. The model fit was excellent, χ^2^(6) = 9.42, *p* = .151, RMSEA=0.026 [0.000, 0.056], CFI=0.996, SRMR=0.018. Regarding covariates, girls had longer total sleep time, earlier sleep onset timing, and faster epigenetic pace of aging. Living closer to the poverty line during childhood was associated with later sleep timing and a faster pace of aging in adolescence. Greater estimated buccal epithelial cell type proportion was associated with a faster pace of aging and accelerated epigenetic clock. Regarding the primary mediation associations, higher levels of childhood home threat were associated with less total sleep time, lower sleep maintenance efficiency, and later sleep onset timing. Lower sleep efficiency and later sleep onset timing were associated with a faster pace of aging and pediatric epigenetic age acceleration. Total sleep time was not associated with epigenetic aging. The total indirect effect of childhood home threat on adolescent epigenetic aging through sleep efficiency and sleep timing was significant for Dunedin pace of aging and PedBE accelerated epigenetic aging. Decomposition of the total indirect effects suggested that the cumulative mediated effect of both sleep efficiency and timing was necessary for PedBE epigenetic age acceleration. The only specific indirect effect that was significant was for later sleep onset timing. Overall, findings support sleep efficiency and onset timing mediating the effects of childhood home threat on adolescent epigenetic aging.

**Table 2.** Primary Path Analysis Model Results.

|  | b | B | B 95% CI | p |
| --- | --- | --- | --- | --- |
| DV: Total Sleep Time |  |  |  |  |
| Home Threat | -.13 | -.06 | -.13 – .00 | .056 |
| Poverty Ratio | -.02 | -.05 | -.10 – .01 | .105 |
| Sex | .40 | .22 | .15 – .28 | < .001 |
| DV: Sleep Efficiency |  |  |  |  |
| Home Threat | -.46 | -.07 | -.14 – .00 | .049 |
| Poverty Ratio | .07 | .06 | -.01 – .12 | .061 |
| Sex | .27 | .05 | -.02 – .11 | .189 |
| DV: Sleep Onset Timing |  |  |  |  |
| Home Threat | .40 | .10 | .04 – .17 | .003 |
| Poverty Ratio | -.06 | -.09 | -.15 – -.04 | .001 |
| Sex | -.44 | -.13 | -.20 – -.06 | < .001 |
| DV: DunedinPACE |  |  |  |  |
| Home Threat | .01 | .04 | -.01 – .08 | .109 |
| Poverty Ratio | -.01 | -.09 | -.13 – -.05 | < .001 |
| Sex | .05 | .14 | .09 – .19 | < .001 |
| Buccal epithelial cell Proportion | .80 | .77 | .75 – .80 | < .001 |
| TST | -.01 | -.01 | -.06 – .05 | .797 |
| Sleep Maintenance Efficiency | -.01 | -.11 | -.16 – -.06 | < .001 |
| Sleep Onset Timing | .01 | .08 | .03 – .12 | .001 |
| DV: PedBE Epigenetic Age Acceleration |  |  |  |  |
| Home Threat | .13 | .07 | .01 – .14 | .042 |
| Poverty Ratio | -.01 | -.04 | -.09 – .02 | .242 |
| Sex | .06 | .04 | -.03 – .10 | .299 |
| Buccal epithelial cell Proportion | 2.49 | .48 | .41 – .55 | < .001 |
| TST | -.05 | -.05 | -.12 – .02 | .175 |
| Sleep Maintenance Efficiency | -.03 | -.09 | -.15 – -.02 | .011 |
| Sleep Onset Timing | .03 | .07 | .01 – .13 | .048 |
| Covariances |  |  |  |  |
| Home Threat WITH Poverty Ratio | -.13 | -.12 | -.18 – -.06 | < .001 |
| TST WITH Efficiency | .24 | .09 | .02 – .16 | .009 |
| TST WITH Sleep Onset Timing | -.16 | -.11 | -.18 – -.03 | .004 |
| Efficiency WITH Sleep Onset Timing | .06 | .01 | -.06 – .08 | .735 |
| DunedinPACE WITH PedBE EAA | .01 | .09 | .01 – .18 | .032 |
| Indirect Effects |  |  |  |  |
| DunedinPACE IND Home Threat | .01 | .02 | .01 – .03 | .006 |
| PedBE IND Home Threat | .03 | .01 | .01 – .02 | .030 |

Note. Model fit: $\chi^2(6) = 9.42$ , $p = .151$ , RMSEA=0.026 [0.000, 0.056], CFI=0.996, SRMR=0.018.

**Figure 3.**
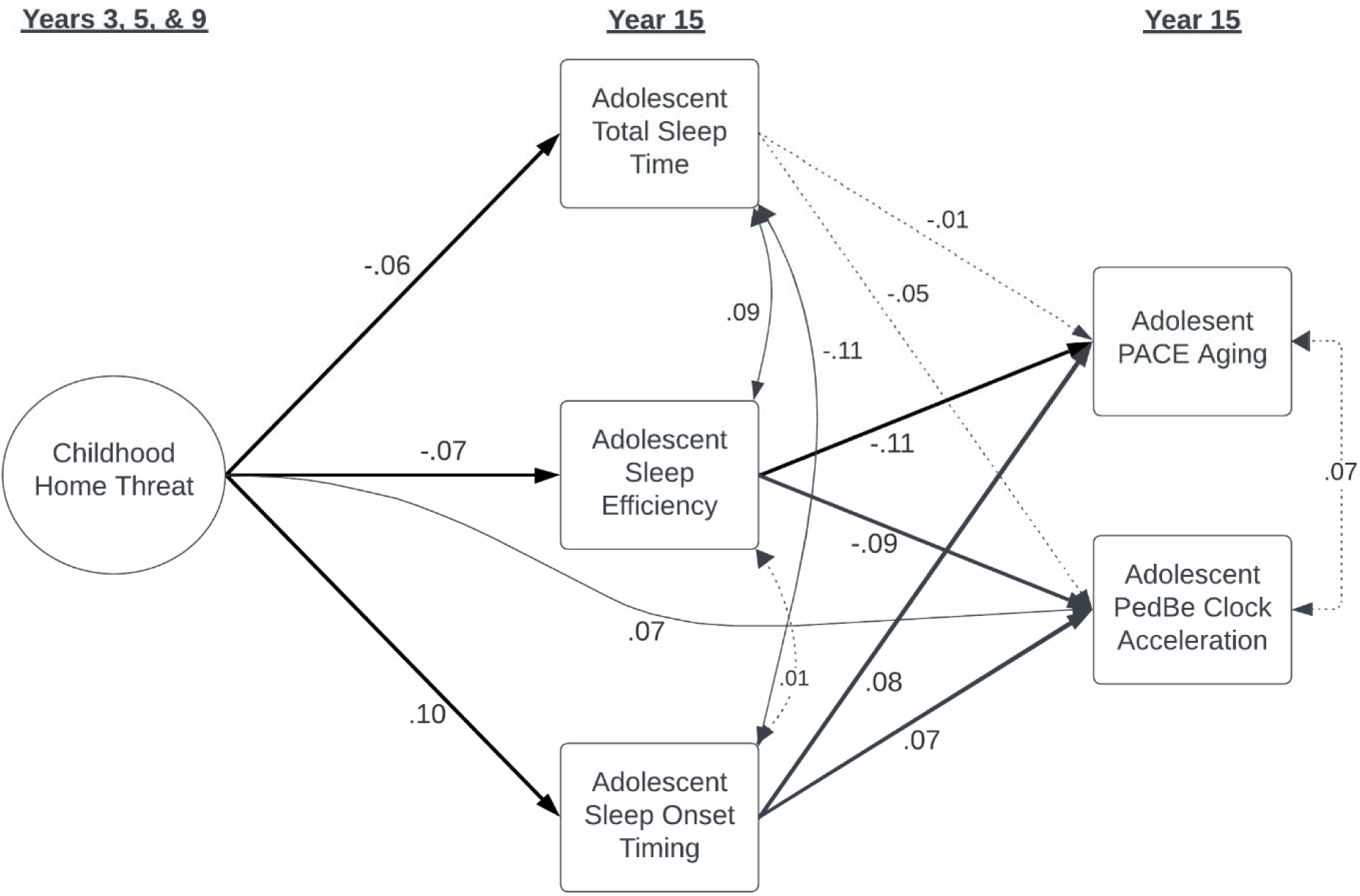
Simplified Path Analysis Model Results. *Note.* Child sex at birth, family poverty ratio at year 1, and BEC proportion are included as covariates in the model but are not depicted above. Solid lines are p < .05, and dotted lines are p ≥ .06. Standardized beta coefficients are depicted. The complete results are in Table 2.

### Sensitivity Analysis

Though details on whether the actigraphy assessment happened during a week when adolescents were attending regular school hours were not available, we created a conservative summer assessment variable based on whether the actigraphy assessment occurred in July or August. Adolescents who did the actigraphy assessment during summer had longer total sleep time, lower sleep efficiency, and later sleep onset timing. When summer assessment was introduced as a covariate into the primary model, associations between home threat and sleep health parameters were similar to the uncorrected model except for sleep efficiency, which was attenuated to *p* = .064 due to increased standard errors. Summer actigraphy timing was not associated with epigenetic outcomes. Next, we examined if differential effects were found when using nighttime sleep duration compared to total 24-hour sleep time (including naps). The strength of the association between higher home threat and shorter nighttime sleep duration (b = -.27, p < .001) increased, but sleep duration remained not associated with epigenetic outcomes. Further, for all sleep health variables we explored possible quadratic effects and found no support for adding nonlinear effects to models.

For primary models, we included participants if they had at least two valid days of actigraphy. For sensitivity analyses, we added the number of valid days into the model as a covariate. For those with two or more days, the number of valid days was not associated with total sleep time, sleep maintenance efficiency, or sleep onset time, and all primary models associated remained unchanged with the inclusion of the number of valid days as a covariate. Finally, given the number of covariates and multiple mediators and outcomes, we applied the Benjamini-Hochberg method to adjust for multiple tests. We found that all p values < .04 had a high confidence of FDR .05 whereas p values ≤ .05 had a moderate confidence of FDR .10. Thus, home threat was a high confidence predictor of later sleep timing but only a moderate confidence predictor of sleep efficiency and duration. Further, sleep efficiency and later sleep onset timing were high-confidence predictors of DunedinPACE, whereas only sleep efficiency was a high-confidence predictor of PedBE epigenetic age acceleration.

## Discussion

The current study used data from a longitudinal birth cohort to evaluate whether three domains of sleep health (i.e., sleep duration, efficiency, and timing) mediated the association between childhood home threat exposure (i.e., physical and emotional abuse) and adolescent epigenetic age acceleration. Overall, findings indicate that adolescent sleep efficiency and sleep onset timing mediated the relationship between childhood home threat exposure and epigenetic aging acceleration.

Findings from the current study provide support for the detrimental impact of early adversity characterized by home-based threat exposure on adolescent sleep health. These results are consistent with prior work showing that childhood adversity is associated with increased subjectively assessed sleep disturbances in FFCWS^17^ and other longitudinal cohorts,^47–49^ as well as actigraphy-assessed sleep duration, efficiency, and timing.^50^ Further, findings showing home-based threat exposure negatively impacting youth sleep are consistent with previous work showing that negative or hostile (e.g., yelling, physical punishment) parenting behaviors are longitudinally associated with subjectively-assessed youth sleep problems (e.g., Acosta et al., 2021) and actigraphy-assessed sleep efficiency.^51^ Overall, our findings are consistent with the sleep-related hyperarousal theoretical model of how home-based adversity may heighten physiological arousal or make it challenging to downregulate physiological stress, resulting in later sleep timing, lower sleep quality, and less total sleep time.^20,52^ Future research will benefit from multiple waves of home-based adversity or stress, physiological pre-sleep arousal (e.g., HR activity), and actigraphy-based sleep health to better disentangle mechanisms or examine bidirectional effects.

To our knowledge, this is the first study to provide evidence that multiple aspects of actigraphy-assessed sleep are associated with accelerated epigenetic aging in adolescence. We found that later sleep timing and lower sleep efficiency were associated with greater epigenetic age acceleration and a faster pace of aging. Though no previous studies have examined these links in adolescence, Larson and colleagues^33^ found that objectively assessed sleep onset timing in adolescents was associated with young adult DNAm in genes previously identified in adult GWAS of sleep and circadian phenotype. Our findings on sleep timing and quality suggest examining sleep health domains beyond sleep duration may be vital in exploring the connection between sleep and epigenetic changes.

Importantly, the current study did not find support for an association between total sleep time and accelerated epigenetic aging. This null finding is inconsistent with Carskadon and colleagues,^29^ who found that short and irregular sleep, assessed via daily diary among emerging adults, may be associated with accelerated epigenetic aging. Further, Koopman-Verhoeff et al.^31^ found one DNAm module associated with actigraphy-assessed sleep duration but no support for sleep timing (assessed by sleep midpoint), though they did not examine epigenetic age acceleration. However, null results for total sleep time in the context of significant results for later sleep timing are consistent with the only other study to examine actigraphy-assessed sleep and epigenetic age acceleration in adolescence.^34^ Banker and colleagues^34^ examined clusters based on sleep duration, sleep timing, and physical activity. They found support for later sleep timing (combined with low physical activity) being associated with more accelerated epigenetic aging in Mexican Adolescents. The current findings, combined with those of Banker and colleagues, support sleep timing as important to consider for understanding how sleep impacts biological embedding via epigenetic changes. Future research will benefit from multiple waves of actigraphy and DNAm along with a more extended period of sleep assessment to explore potential mechanisms or alternative explanations, such as sleep irregularity,^29^ sleep debt,^53,54^ social jetlag,^17^ chronotype,^55^ disruption of circadian-related physiological systems (e.g., HPA-axis^20^), or misalignment between central and peripheral clocks.^56^

The current study was the first to examine sleep disruption as a mechanism linking early childhood adversity and adolescent biological embedding via epigenetic age acceleration. Findings in the current study supported sleep timing and quality as mediators for the longitudinal association between home threat exposure and Dunedin pace of aging and pediatric epigenetic age acceleration, with the greatest support for sleep efficiency or the combined mediation effect of both sleep health parameters. Further, findings were robust to controlling for family poverty level, child sex, timing of actigraphy assessment, and adjusting for multiple tests. Though limited by sleep and epigenetic aging being assessed at the same wave (Year 15), these results provide preliminary support for sleep as a mechanism for adversity-related biological embedding and long-term health trajectories. Findings warrant greater attention to pediatric sleep health in theoretical models of biological embedding of adversity and point to future directions in improving sleep health to potentially prevent adversity-related epigenetic age acceleration.

The current study has notable strengths and some limitations that would benefit from future research. This study featured a longitudinal design that allowed the exploration of directions of associations, which has been a limitation in past research. However, the single wave of actigraphy-based sleep assessment limited our ability to examine the bidirectional or alternative direction of effects between adversity and sleep and between sleep and epigenetic outcomes. Thus, actigraphy-based sleep before home threat exposure and three longitudinal waves of both sleep and DNAm are needed for more robust causal inferences. Alternatively, future research could benefit from experimental designs that improve sleep health and examine downstream impacts on epigenetic age acceleration. In the area of parenting, emerging evidence suggests enhancing family health and parent-child interaction may have a preventative effect on epigenetic age acceleration^57,58^ or pace of aging^59^ among youth with heightened levels of adversity. Thus, future studies could explore how experimentally enhancing pediatric sleep health could result in reduced epigenetic age acceleration.

Another strength is its multi-modal approach, which utilizes parent-report, actigraphy-assessed sleep health, and DNAm-based biomarkers of aging, therefore increasing confidence in associations observed due to removing potential method-effect confounding. Additionally, the current study included a large racially and economically diverse sample of youth across the United States. Research on social epigenomics has predominately focused on White European-ancestry adults. In contrast, social epigenetic research involving US racial and ethnic minority populations and other populations experiencing health disparities (e.g., low socioeconomic status) remained limited, substantially limiting advancements being applied toward understanding or eliminating health disparities.^60^ Findings from the current study suggest potential future research directions for connecting largely separate literatures on the biological embedding of health disparities via epigenetic changes ^60^and the causes and consequences of sleep health disparities.^61,62^ Research on family or community-level intervention or prevention programs that simultaneously support family health (e.g., parenting, parental well-being) and adolescent sleep health may be particularly advantageous future directions for potentially preventing accelerated epigenetic aging.

## Data Availability Statement

The data underlying this article is publicly available and can be accessed via https://ffcws.princeton.edu/. Complete code and output for final models will be made available on an Open Science Framework page and linked to the final publication.

